# Shift work is associated with positive COVID-19 status in hospitalised patients

**DOI:** 10.1101/2020.12.04.20244020

**Authors:** Robert Maidstone, Simon G Anderson, David W Ray, Martin K Rutter, Hannah J Durrington, John F Blaikley

## Abstract

**Introduction:** Shift work is associated with both mental, and physical ill health, including lung disease and infections. However, the impact of shift work on significant COVID-19 illness has not be assessed. We therefore investigated whether shift work is associated with COVID-19.

**Methods:** 501,000 UK biobank participants were linked to secondary care SARS-CoV-2 PCR results from public health England. Healthcare workers and those without an occupational history were excluded from analysis.

**Results:** Multivariate logistic regression taking into account age, sex, ethnicity and deprivation index revealed that irregular shift work (OR 2.42 95%CI 1.92-3.05), permanent shift work (OR 2.5, 95%CI 1.95-3.19), day shift work (OR 2.01, 95%CI 1.55-2.6), irregular night shift work (OR 3.04, 95%CI 2.37-3.9) and permanent night shift work (OR 2.49, 95%CI 1.67-3.7) were all associated with positive COVID-19 tests compared to participants that did not perform shift work. This relationship persisted after adding sleep duration, chronotype, pre-morbid disease, BMI, alcohol and smoking. Work factors (proximity to a colleague combined with estimated disease exposure) were positively correlated with COVID-19 incidence (r^2^=0.248, p=0.02). If this was added to the model shift work frequency remained significantly associated with COVID-19. To control for non-measured occupational factors the incidence of COVID-19 in shift workers was compared to colleagues in the same job who did not do shift work. Shift workers had a higher incidence of COVID-19 (p<0.01).

**Conclusions:** Shift work is associated with a higher likelihood of in-hospital COVID-19 positivity. This risk could potentially be mitigated via additional workplace precautions or vaccination.

## Introduction

The COVID-19 pandemic has affected millions of people so far. There are limited therapeutic options for COVID-19 causing management to focus on containment^1^. A greater understanding of risk factors for COVID-19 susceptibility permits protection of the most vulnerable, mitigates occupational exposure and allows for more effective targeting of vaccines^2 3^. Several risk factors have already been identified for COVID-19 including age, obesity, sex, ethnicity, and comorbidities^2 3^. Occupation has also been recognised as a risk factor for COVID-19 infection with healthcare workers in patient-facing roles being at highest risk^4-6^. However, the type of working patterns have not been extensively studied despite COVID-19 outbreaks occurring at food-processing factories where nightshift workers were employed^7^.

Worldwide shift work is becoming increasingly common with 10-40% of workers in most countries being involved^8^. The adverse health effects of shift work are increasingly being recognised. Shift work is associated with respiratory disease^9 10^, diabetes^11^, cancer^12^, and non-COVID-19 infectious diseases^13 14^. The mechanisms underlying these associations remain uncertain, however, sleep disruption, poor diet and circadian misalignment may account for some of the effects^15^.

As the immune system is regulated by the circadian clock, it has been hypothesised that shift work-induced circadian misalignment could increase susceptibility to COVID-19 infection^16^. Current UK guidance from the Health and Safety executive advocates shift working where possible, to limit the number of people in the workplace at any one time^17^. Therefore, we aimed to investigate the association between shift work status and COVID-19 infection, using the UK Biobank^18^.

## Methods

The UK Biobank study was approved by the National Health Service National Research Ethics Service (ref. 11/NW/0382) and HTA (IRAS 282966). All participants provided written informed consent to participate in the UK Biobank.

### Participants

We studied UK Biobank participants after excluding the following groups: a) healthcare workers, on the basis that their occupation puts them at an especially increased risk of COVID-19 infection, and they have a high prevalence of shift working; b) participants who had COVID-19 testing outside of secondary care; and c) people who had not provided a detailed job history to determine shift work status.

### Shift work frequency assessment

Shift work was defined as previously reported^9^. Briefly, participants employed at baseline between 2006 and 2010 were asked to report whether their current main job involved shift work (i.e. a schedule falling outside of 9:00am to 5:00pm). Such schedules involved afternoon, evening or night shifts (or rotating though these shifts). Participants could respond ‘never/rarely’, ‘sometimes’, ‘usually’, ‘always’, ‘prefer not to answer’ and ‘do not know’. For analysis in this study those that answered ‘never/rarely’ were defined as never, those that answered ‘sometimes’ or ‘usually’ were defined as irregular shift workers, and those that answered ‘always’ were defined as permanent. If participants recorded the additional options of ‘prefer not to answer’ or ‘do not know’, they were excluded from shift work frequency analysis.

### Shift work type assessment

All participants except those that ‘never’ performed shift work were included in shift work type analysis. They were then asked whether their main job involved night shifts, defined as ‘a work schedule that involves working though the normal sleeping hours, for instance, working though the hours from 12:00am to 6:00am’. Response options were ‘never/rarely’, ‘sometimes’, ‘usually’, or ‘always’ and included additional options: ‘prefer not to answer’ and ‘do not know’. Based on these responses and whether they did shift work, we derived participants’ type of shift work, categorized as, ‘none’ (work between hours 9am-5pm), ‘day shift’, ‘irregular night shift work’ (those who answered sometimes or usually) and ‘permanent night shift work’. Participants responding ‘prefer not to answer’ or ‘do not know’, were excluded from this analysis.

### COVID-19 positive case definition

Cases of COVID-19 were defined by a positive PCR for Sars-CoV2 from nasopharyngeal swabs taken from the 16 March to the 24 August 2020 and recorded by public health England(PHE)^19^. We confined analysis to those people with an in-hospital PCR test.

### Chronotype

Participants self-reported chronotype on a touch-screen questionnaire at baseline by answering the question: “Do you consider yourself to be….” with response options “Definitely a ‘morning’ person”, “More a ‘morning’ than ‘evening’ person”, “More an ‘evening’ than a ‘morning’ person,” “Definitely an ‘evening’ person,” “Do not know,” and “Prefer not to answer.” Subjects who responded “Do not know” or “Prefer not to answer” were set to missing. This single item has been shown to correlate with sleep timing and dim-light melatonin onset. For our analyses we combined “more a ‘morning’ than ‘evening’ person” with “more an ‘evening’ than ‘morning’ person” to form an intermediate group.

### Occupation ‘Proximity Score’

The average physical distance between two individuals employed in particular occupations has been estimated by the ‘Proximity Score’. We obtained these scores from the Office for National Statistics(ONS)^6^ O*NET database based on workers responses to a question “how physically close to other people are you when you perform your current job?”. The answer was then scaled out of 100 and mapped onto the four-digit Standard Occupational Classification (SOC) available for UK Biobank participants.

### Occupation ‘Exposure Score’

The Exposure Score is a measure of the exposure of an individual to a disease^6^. We also obtained these data from the ONS^6^ based on responses to a question “How often does your current job require that you be exposed to diseases or infection?”. The answer was scaled out of 100 and mapped onto a four-digit SOC in UK Biobank participants.

### Work Environment Score

The Work Environment Score was defined as the sum of the Proximity and Exposure scores.

### Statistical Analysis

We employed a multivariate logistic regression model to the data and used this to estimate adjusted odds ratios and 95% asymptotic confidence intervals on those odds ratios. Covariates were defined using data collected at the time of enrolment into the UK Biobank. In model 1 we initially adjusted for age, sex, ethnicity and Townsend Deprivation Index (TDI). We extend this adjustment in model 2 to additionally include sleep duration. Lastly, model 3 also included smoking history, alcohol history, BMI, hypertension, diabetes, chronotype, cardiovascular disease, renal failure, liver disease, asthma and COPD. An ANOVA was used when investigating continuous variables and a Chi squared test for categorical variables. R (v4.0.2) was used to analyse data. R packages used include; flex table (v0.5.11), Magritte (v1.5), officer (v0.3.14) and tidy verse (v1.3.0).

### Patient/ Public involvement

Participants were not involved in the design or analysis of this study.

### Sensitivity analysis

In participants with proximity and exposure data(n=167,318), we undertook sensitivity analyses to account for the addition of *work environment scores* into model 3, by performing additional analyses after further adjustment for this covariate.

Since the work environment score may not fully reflect all the work environment risk factors we compared COVID-19 positivity in shift workers and non-shift workers who shared the same job type (SOC code) when there was at least 1 positive COVID case per job type.

## Results

### Demographics

The UK Biobank included 502,450 participants from which we excluded 1,086 healthcare workers, and 3,050 participants who had COVID-19 testing outside of secondary care (**Suppl. Fig. 1**). For frequency of shift work analysis 214,377 participants were excluded since they were not in full time employment or declined to answer leaving 284,027 participants. Of these standard occupational classification (SOC) job codes could be matched to 197,790 participants. For type of shift work analysis 214,035 participants were excluded since they were not in full time employment or declined to answer leaving 284,629 participants. Of these SOC job codes could be matched to 198,061 participants (**Suppl. Fig. 1**).

### Clinical characteristics

Clinical characteristics are shown in table 1 for shift work frequency and supplement table 1 for type of shift work. Shift workers tended to be younger, male, have a higher BMI, smoke more, have a lower alcohol intake, non-White ethnicity, and higher levels of deprivation. Furthermore, they were more likely to have comorbid disease.

**Table 1:** Shift work frequency: Demographics by current shift work exposure (N = 284,027): Variables are expressed as mean (±SD) or as percentages

|  | Reported Shift work frequency |  |  | P Values |
| --- | --- | --- | --- | --- |
|  | Never Shift Workers | Irregular shift work | Permanent shift work |  |
| N | 235135 | 27056 | 21836 |  |
| Age (years) | 52.9 (7.12) | 52.16 (7.09) | 51.44 (6.86) | <0.01 |
| Sex (% male) | 46.61 | 54.83 | 55.83 | <0.01 |
| BMI (kg/m<sup>2</sup>) | 27.09 (4.65) | 27.91 (4.92) | 28.23 (4.98) | <0.01 |
| **Smoker (%)** |  |  |  | <0.01 |
| Never | 58.11 | 53.09 | 52.97 |  |
| Previous | 31.89 | 31.71 | 30.77 |  |
| Current | 9.75 | 14.77 | 15.92 |  |
| Smoking pack-years | 19.99 (16) | 23.59 (17.97) | 24.04 (17.51) | <0.01 |
| Daily alcohol intake (%) | 20.46 | 17.81 | 12.89 | <0.01 |
| Sleep Duration (h) | 7.05 (1.03) | 6.92 (1.21) | 6.81 (1.39) | <0.01 |
| **Chronotype (%)** |  |  |  | <0.01 |
| Morning | 23.34 | 24.51 | 22.55 |  |
| Evening | 8.01 | 9.04 | 10.97 |  |
| **Ethnicity (%)** |  |  |  | <0.01 |
| White British | 88.5 | 82.06 | 81.77 |  |
| White Other | 6.44 | 7.27 | 6.41 |  |
| Mixed | 0.65 | 0.93 | 0.89 |  |
| Asian | 1.71 | 3.63 | 3.54 |  |
| Black | 1.39 | 3.26 | 4.6 |  |
| Chinese | 0.34 | 0.62 | 0.32 |  |
| Other | 0.69 | 1.85 | 2.11 |  |
| Weekly work hours | 34.24 (13.19) | 37.05 (14.77) | 37.68 (12.55) | <0.01 |
| Single Occupancy (%) | 15.63 | 18.49 | 18.99 | <0.01 |
| Urban area (%) | 85.98 | 89.1 | 90.42 | <0.01 |
| Townsend Index | -2.24 (-3.7 to 0.18) | -1.43 (-3.25 to 1.55) | -1.05 (-3.02 to 1.95) | <0.01 |
| High Cholesterol (%) | 7.88 | 8.48 | 8.89 | <0.01 |
| Diabetes (%) | 3.22 | 4.35 | 4.58 | <0.01 |
| Hypertension (%) | 20.33 | 22.25 | 22.46 | <0.01 |
| Depression (%) | 4.61 | 4.84 | 5.21 | <0.01 |
| Cardiovascular Disease (%) | 2.27 | 2.74 | 2.54 | <0.01 |
| Impaired Renal Function (%) | 0.09 | 0.11 | 0.11 | 0.52 |
| Defined Asthma (%) | 4.93 | 5.05 | 5.12 | 0.32 |
| COPD (%) | 0.13 | 0.2 | 0.2 | <0.01 |
| Liver Disease (%) | 0.53 | 0.55 | 0.47 | 0.58 |

Within the UK Biobank 6,442 participants had in-hospital COVID-19 testing, with 498 testing positive. Of these, 316 did not work shifts (‘never’ only worked between 9am-5pm), 98 worked irregular shifts and 84 worked permanent shifts.

### Association between shift work frequency and COVID-19

To ascertain whether shift work is associated with in hospital COVID-19 positive test we compared workers who never worked shifts with participants who worked irregular or permanent shifts. Shift work was associated with a higher likelihood of COVID-19 for both irregular (OR 2.42 (95% CI 1.92-3.05)) and permanent shift work (OR 2.50 (1.95-3.19)) after adjusting for age, sex, ethnicity, and Townsend Deprivation Index (TDI) (model 1, **Fig 1a**). One of the characteristic features of shift work is sleep disruption, and in particular sleep deprivation. After adjustment for sleep duration the odds ratios remained broadly unchanged (model 2, **Fig. 1a**). As shift work is associated with obesity, smoking, alcohol intake, and as chronotype impacts on nightshift tolerability we adjusted for BMI, chronotype, alcohol intake, smoking and prior disease (model 3). The association with irregular shift work remained (OR 2.29 (1.53-3.45) following this adjustment and increased for permanent shift work OR 2.68 (1.78-4.03)) (model 3, **Fig. 1a**).

**Figure (1):**
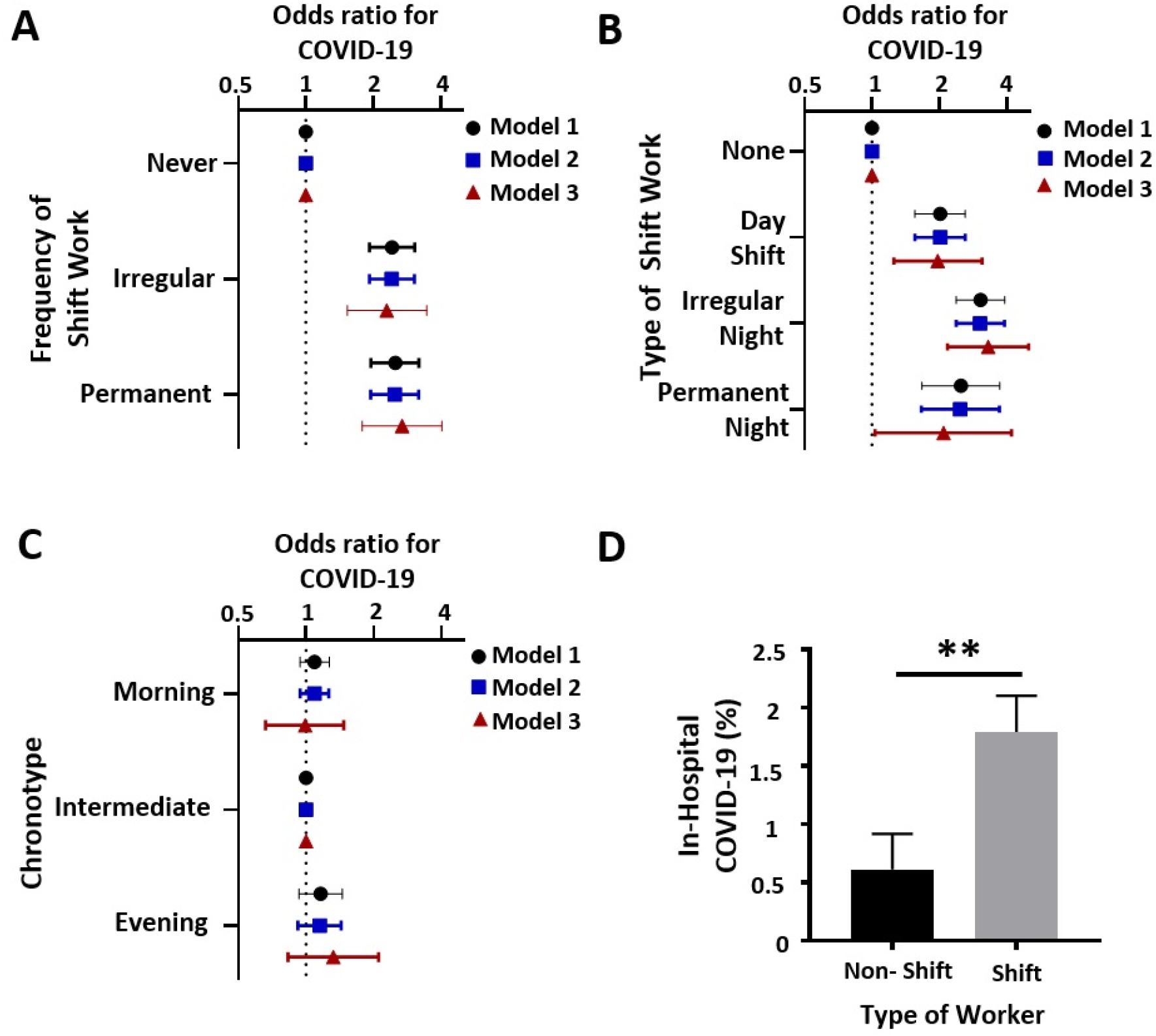
Shift work is associated with COVID-19: Workers were stratified by work pattern in the UK biobank. **A** shows the association of shift work frequency with COVID-19. **B** shows the association of shift work type with COVID-19. **C** shows the association of chronotype with COVID-19. Model 1 adjusts for the covariates age, sex, Townsend Deprivation Index (TDI) and ethnicity. Model 2 extended the adjustment to include sleep duration. Model 3 also includes smoking history, alcohol history, BMI, hypertension, diabetes, cardiovascular disease, renal failure, liver disease, asthma and COPD. Chronotype was also included in model 3 for **A** and **B. D** shows the difference in COVID-19 frequency between shift workers and non-shift workers who do the same job according to SOC code (n=38 jobs). Forrest plots of odds ratios (ORs) for COVID-19 with 95% confidence intervals are shown. **=p<0.01 paired t test (mean±SEM)

### Association between type of shift work and COVID-19

Next, we investigated whether the type of shift work affected the association with COVID-19. Compared to workers who engaged in no shift work (‘none’), day shift workers and night shift workers (working irregular and permanent night shifts) had a higher likelihood of having a positive COVID-19 test after adjustment for age, sex, ethnicity and TDI. (**Fig. 1b**, model 1). In the same model, irregular night shift work was associated with a higher likelihood of having COVID-19 during hospitalisation (OR 3.04 (2.37-3.90)), and permanent night shift work was also associated with higher odds (OR 2.49 (1.67-3.70)). Surprisingly, we also found that workers who worked day shifts also had a higher likelihood of COVID-19 (OR 2.01 (1.55-2.60)) compared to those reporting no history of shift work. After adjusting for sleep duration the odds ratios remained largely unchanged (**Fig. 1b**, model 2). Analysis using Model 3 also showed a positive association between irregular night shift work and COVID-19 (OR 3.29 (2.17-4.98)), for permanent night shift workers (OR of 2.08 (1.03-4.18)) and for day shift workers (OR of 1.96 (1.25-3.09)), **Fig. 1b**.

### Chronotype and COVID-19

One possible mechanism for the effects of shift work is through circadian misalignment^9^. Individuals with extreme chronotypes live misaligned even when not shift working. We found no chronotype association with COVID-19 (**Fig. 1c**).

### Job characteristics and COVID-19

COVID-19 risk is associated with job type^4-6^, possibly mediated via proximity to other workers or exposure to the disease^6^. There was no correlation between ‘*Proximity Score’* and COVID-19 positive tests (r^2^=-0.166, p=0.98; **Suppl. Fig. 2A**) or between ‘*Exposure Score’* and COVID-19 positive tests (r^2^=0.2386, p=0.09; **Suppl. Fig. 2B**). However, there was a positive correlation between work environment score (combined exposure and proximity score) and COVID-19 (**Supplement Fig. 2C**, r^2^=0.248, p=0.02).

### Sensitivity analyses

Exposure, proximity and work environment scores were all higher in shift workers (day n=15,442 and night n=15,610) compared to non-shift workers (‘none’, n=168,617) (**Suppl. Table 2**), suggesting that the type of job may differ between non-shift workers and shift workers. Therefore, we undertook sensitivity analyses to account for the addition of *work environment scores* in model 3.

For frequency of shift work, after adjusting for model 3 covariates and *work environment* both irregular shift workers (n=17,880, OR 1.95 (1.12-3.39)) and permanent shift workers (n=12,592, OR

2.01 (1.1-3.69)) had a higher likelihood of COVID-19 when compared to never shift workers (n=167,318) (**Suppl. Fig. 2D**) When type of shift work was examined, after adjusting for the same covariates, compared to non-shift workers there was an association between irregular night shift work (n= 11,173 OR 2.59 (1.45-4.62)) and COVID-19 (**Suppl. Fig. 2E**). However, no significant association for day shift workers (n=15,267, OR 1.74 (0.92-3.26) or permanent night shift workers (n=4,303, OR 1.13 (0.35-3.68)) was found; possibly because of type 2 errors.

We compared COVID-19 positivity in shift workers and non-shift workers who shared the same job type (SOC code). Shift workers had a higher rate of COVID-19 compared to non-shift workers (n=38 jobs) (**Fig 1d)**.

## Discussion

We now show that shift workers have higher odds of testing positive for COVID19 in hospital compared to non-shift workers. Both permanent and irregular shift workers (encompassing both day and night shift workers) had increased odds, compared to workers who never worked shifts. When we stratified shift workers into day shift and night shift workers (including permanent and irregular night shifts), we found that the association with COVID-19 hospitalisation remained increased regardless of the time of day of shift. Sensitivity analysis further revealed that in a sub-group of participants a combination of proximity and exposure scores for job type did not, explain the association between shift work and COVID-19 positivity. As healthcare workers frequently work shifts and are exposed to higher risks of COVID-19 infection by type of work we excluded them from our analysis.

The size of effect of shift work as a risk factor for COVID-19 is comparable to other reported risk factors for COVID-19 such as being non-white, being most socioeconomically deprived, and having a BMI ≥40 kg/m2^3^. Strikingly, compared to the odds ratios reported for shift work effects in other diseases in the UK Biobank, in this study the effects of shift work were much bigger, suggesting this is an important risk factor should be considered in future public health measures. A key difference with shift work compared to most other COVID-19 risk factors is that this risk could be mitigated relatively quickly. Possible solutions are increasing distance between workers, wearing personal protective equipment and enhanced cleaning of the workspace.

One potential explanation for the effect of shift work on COVID-19 hospitalisation is through the mechanism of circadian misalignment. Supporting this hypothesis is the discovery that melatonin, a drug which can entrain circadian rhythmicity, is protective against COVID-19^20^. Early chronotypes experience circadian misalignment when working night shifts and find it difficult to adjust, whereas late chronotypes experience similar disruption when working early shifts^21^. Therefore, we determined if there was an association between chronotype and COVID-19 hospitalisation. However, no such association was observed. The low numbers of COVID-19 cases for each extreme chronotype (n= 274 morning, 94 evening) suggest that this study may have been underpowered to detect a significant difference for a modest effect comparable to the effect sizes for chronotype in other UK biobank studies. Repeating this analysis would be helpful if COVID-19 cases continue to rise.

Another possible explanation for our results is that the type of jobs done by shift workers might increase the association with COVID-19. We did this in three ways, firstly by excluding healthcare workers *a priori* from analysis. Secondly, we used data from the Office for National Statistics (ONS) regarding worker proximity and disease exposure and were able to match these codes to 2/3rds of the occupations listed for UK biobank participants. After accounting for worker proximity and disease exposure, statistical significance was lost for some exposures, but the strength and direction of effect remained. We believe these observations are explained by reduced power since some of the categories had only 20 positive COVID cases. Thirdly, we performed an intra-job comparison between shift workers and those that did not perform shift work which showed higher rates of COVID-19 in the shift work group. Possible explanations for the observed higher rate of COVID-19 might include increased occupancy of workspaces over 24 hours for shift workers, reduced time for cleaning between shifts and tiredness resulting in less awareness of health and safety measures.

Recently shift work has been shown to alter how the immune system responds to infection and several epidemiological studies have identified that shift workers are more prone to infections^13 14^. Shift work was not included in the ISARIC study^22^ and has not been included as a co-variate in other large epidemiological studies^23 24^. The large association reported in this study would suggest that shift work should be included in future epidemiological pandemic protocols, especially since shift work has been linked to a number of health conditions^25^ including diabetes, obesity, cancer, fibrosis and asthma that altered COVID-19 risk for this pandemic.

The strengths of this study are the large number of individuals >280,000 participants that were analysed. Participants were also recruited before the pandemic permitting the control, i.e. non COVID-19, group to be selected without bias. However, there are weaknesses in our study. Data collected by questionnaire for the UK Biobank and used in this study was recorded ten years before COVID-19 and although some of the data has been updated through Hospital episode statistics it cannot be viewed as a contemporaneous record. We have also previously shown that in this cohort at the time of data collection the average length time spent in their current job was 20 years regardless of shift work status^9^. Lastly, accounting for collider bias^26^ in analyses on the UK Biobank data is a non-trivial task, and analysis on COVID-19 disease risk is particularly susceptible to this. We hope to have mitigated this by presenting multiple models of differing complexities, as well as a job paired analysis of the effect of shift work (Figure 1D). Despite this, it should still be noted that any conclusions drawn here are made in relation to the UK Biobank cohort only and therefore need to be validated in other populations.

We defined COVID-19 as a positive SARS-COV2 test taking place in secondary care. This approach has previously been validated^19^ and identifies those individuals with a more severe form of COVID-19, although we acknowledge that a minority of our cohort may have been picked up during hospital screening. Focusing our research on a more severe type of COVID-19 is important as it is this group of patients that should be targeted for vaccination or enhanced infection control if COVID-19 associated mortality is to be reduced.

## Conclusion

We show that there is an increased likelihood of COVID-19 in shift workers which is comparable to known COVID-19 risk factors. We would advocate that shift work is treated as a modifiable risk factor for COVID-19. Sensible precautions in the workplace might include increased cleaning schedules, reduced numbers of workers on any one shift, providing personal protective equipment to shift workers and targeting shift workers for early COVID-19 vaccination programmes.

## Data Availability

Data will be made available upon reasonable request when the study has been formally published.

## Contribution

JFB, HD, MKR, DWR designed the study, SA RM and JFB analysed the data, all authors wrote the paper and provided critical analysis. JB and HD accepts full responsibility for the work, had access to the data and controlled the decision to publish. The corresponding authors attest that al listed authors meet authorship criteria and that no others meeting the criteria have been omitted.

## References

1. Wilder-Smith A, Chiew CJ, Lee VJ. Can we contain the COVID-19 outbreak with the same measures as for SARS? The Lancet Infectious diseases 2020;20(5):e102–e07. doi: 10.1016/s1473-3099(20)30129-8 [published Online First: 2020/03/09]

2. Clift AK, Coupland CAC, Keogh RH, et al. Living risk prediction algorithm (QCOVID) for risk of hospital admission and mortality from coronavirus 19 in adults: national derivation and validation cohort study. BMJ (Clinical research ed) 2020;371:m3731. doi: 10.1136/bmj.m3731 [published Online First: 2020/10/22]

3. McQueenie R, Foster HME, Jani BD, et al. Multimorbidity, polypharmacy, and COVID-19 infection within the UK Biobank cohort. PloS one 2020;15(8):e0238091. doi: 10.1371/journal.pone.0238091 [published Online First: 2020/08/21]

4. Nguyen LH, Drew DA, Graham MS, et al. Risk of COVID-19 among front-line health-care workers and the general community: a prospective cohort study. The Lancet Public health 2020;5(9):e475–e83. doi: 10.1016/s2468-2667(20)30164-x [published Online First: 2020/08/04]

5. Mutambudzi M, Niedzwiedz CL, Macdonald EB, et al. Occupation and risk of severe COVID-19: prospective cohort study of 120,075 UK Biobank participants. medRxiv 2020:2020.05.22.20109892. doi: 10.1101/2020.05.22.20109892

6. Statisics OfN. Which occupations have the highest potential exposure to the coronavirus (COVID- 19)? : Office for National Statistics; 2020 [Available from: https://www.ons.gov.uk/employmentandlabourmarket/peopleinwork/employmentandemployeetypes/articles/whichoccupationshavethehighestpotentialexposuretothecoronaviruscovid19/2020-05-11 accessed 7th november 2020.

7. Waltenburg MA, Victoroff T, Rose CE, et al. Update: COVID-19 Among Workers in Meat and Poultry Processing Facilities - United States, April-May 2020. MMWR Morbidity and mortality weekly report 2020;69(27):887–92. doi: 10.15585/mmwr.mm6927e2 [published Online First: 2020/07/10]

8. Mariya Aleksynska JB, David Foden, Hannah Johnston, Agnès Parent-Thirion, Julie Vanderleyden. Working conditions in a global perspective. Geneva: Office of the European Union, Luxembourg, and International Labour Organization, 2019.

9. Maidstone RJ, Turner J, Vetter C, et al. Night shift work is associated with an increased risk of asthma. Thorax 2020 doi: 10.1136/thoraxjnl-2020-215218 [published Online First: 2020/11/18]

10. Cunningham PS, Meijer P, Nazgiewicz A, et al. The circadian clock protein REVERBα inhibits pulmonary fibrosis development. Proceedings of the National Academy of Sciences of the United States of America 2020;117(2):1139–47. doi: 10.1073/pnas.1912109117 [published Online First: 2019/12/28]

11. Vetter C, Dashti HS, Lane JM, et al. Night Shift Work, Genetic Risk, and Type 2 Diabetes in the UK Biobank. Diabetes Care 2018;41(4):762–69. doi: 10.2337/dc17-1933 [published Online First: 2018/02/15]

12. Kervezee L, Shechter A, Boivin DB. Impact of Shift Work on the Circadian Timing System and Health in Women. Sleep medicine clinics 2018;13(3):295–306. doi: 10.1016/j.jsmc.2018.04.003 [published Online First: 2018/08/14]

13. Mohren DC, Jansen NW, Kant IJ, et al. Prevalence of common infections among employees in different work schedules. Journal of occupational and environmental medicine 2002;44(11):1003–11. doi: 10.1097/00043764-200211000-00005 [published Online First: 2002/11/27]

14. Loef B, van Baarle D, van der Beek AJ, et al. Shift Work and Respiratory Infections in Health-Care Workers. American journal of epidemiology 2019;188(3):509–17. doi: 10.1093/aje/kwy258 [published Online First: 2018/11/27]

15. James SM, Honn KA, Gaddameedhi S, et al. Shift Work: Disrupted Circadian Rhythms and Sleep- Implications for Health and Well-Being. Current sleep medicine reports 2017;3(2):104–12. doi: 10.1007/s40675-017-0071-6 [published Online First: 2017/10/24]

16. Lim RK, Wambier CG, Goren A. Are night shift workers at an increased risk for COVID-19? Medical hypotheses 2020;144:110147. doi: 10.1016/j.mehy.2020.110147 [published Online First: 2020/08/08]

17. Social distancing to make your workplace COVID-secure: Health and Safety Executive; 2020 [updated 2nd November. Available from: https://www.hse.gov.uk/coronavirus/social-distancing/arriving-leaving.htm accessed 7th Novemebr 2020.

18. Palmer LJ. UK Biobank: bank on it. Lancet (London, England) 2007;369(9578):1980–82. doi: 10.1016/s0140-6736(07)60924-6 [published Online First: 2007/06/19]

19. Armstrong J, Rudkin JK, Allen N, et al. Dynamic linkage of COVID-19 test results between Public Health England’s Second Generation Surveillance System and UK Biobank. Microbial genomics 2020;6(7) doi: 10.1099/mgen.0.000397 [published Online First: 2020/06/20]

20. Zhou Y, Hou Y, Shen J, et al. A network medicine approach to investigation and population-based validation of disease manifestations and drug repurposing for COVID-19. PLoS biology 2020;18(11):e3000970. doi: 10.1371/journal.pbio.3000970 [published Online First: 2020/11/07]

21. Juda M, Vetter C, Roenneberg T. Chronotype modulates sleep duration, sleep quality, and social jet lag in shift-workers. Journal of biological rhythms 2013;28(2):141–51. doi: 10.1177/0748730412475042 [published Online First: 2013/04/23]

22. Docherty AB, Harrison EM, Green CA, et al. Features of 20?133 UK patients in hospital with covid- 19 using the ISARIC WHO Clinical Characterisation Protocol: prospective observational cohort study. BMJ (Clinical research ed) 2020;369:m1985. doi: 10.1136/bmj.m1985 [published Online First: 2020/05/24]

23. Richardson S, Hirsch JS, Narasimhan M, et al. Presenting Characteristics, Comorbidities, and Outcomes Among 5700 Patients Hospitalized With COVID-19 in the New York City Area. Jama 2020;323(20):2052–59. doi: 10.1001/jama.2020.6775 [published Online First: 2020/04/23]

24. Garg S, Kim L, Whitaker M, et al. Hospitalization Rates and Characteristics of Patients Hospitalized with Laboratory-Confirmed Coronavirus Disease 2019 - COVID-NET, 14 States, March 1-30, 2020. MMWR Morbidity and mortality weekly report 2020;69(15):458–64. doi: 10.15585/mmwr.mm6915e3 [published Online First: 2020/04/17]

25. Kecklund G, Axelsson J. Health consequences of shift work and insufficient sleep. BMJ (Clinical research ed) 2016;355:i5210. doi: 10.1136/bmj.i5210 [published Online First: 2016/11/03]

26. Griffith GJ, Morris TT, Tudball MJ, et al. Collider bias undermines our understanding of COVID-19 disease risk and severity. Nature communications 2020;11(1):5749. doi: 10.1038/s41467-020-19478-2 [published Online First: 2020/11/14]

