## Supplement for "Shift work is associated with positive COVID-19 status in hospitalised patients"

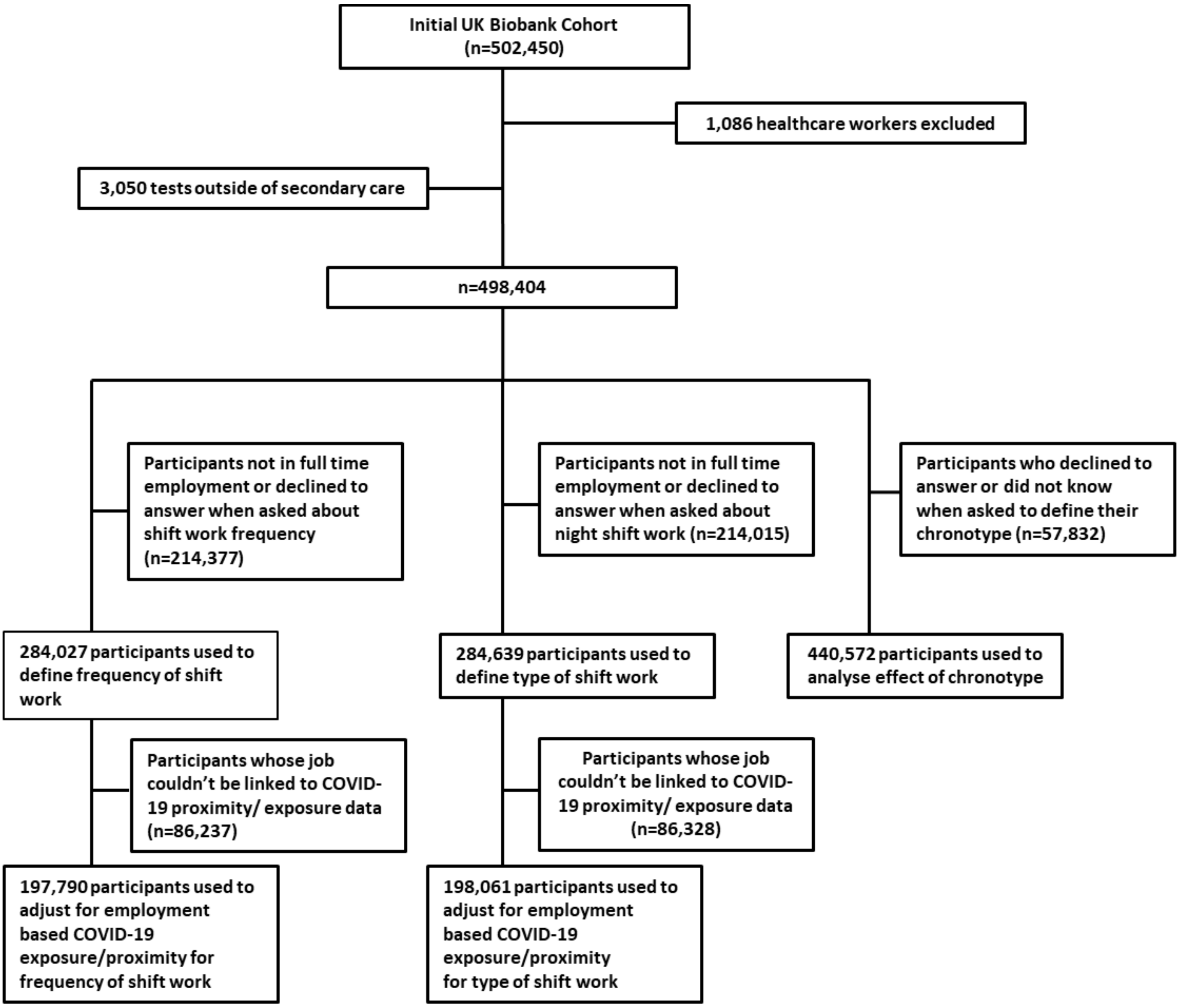


**Suppl. Figure 1: Strobe Diagram**

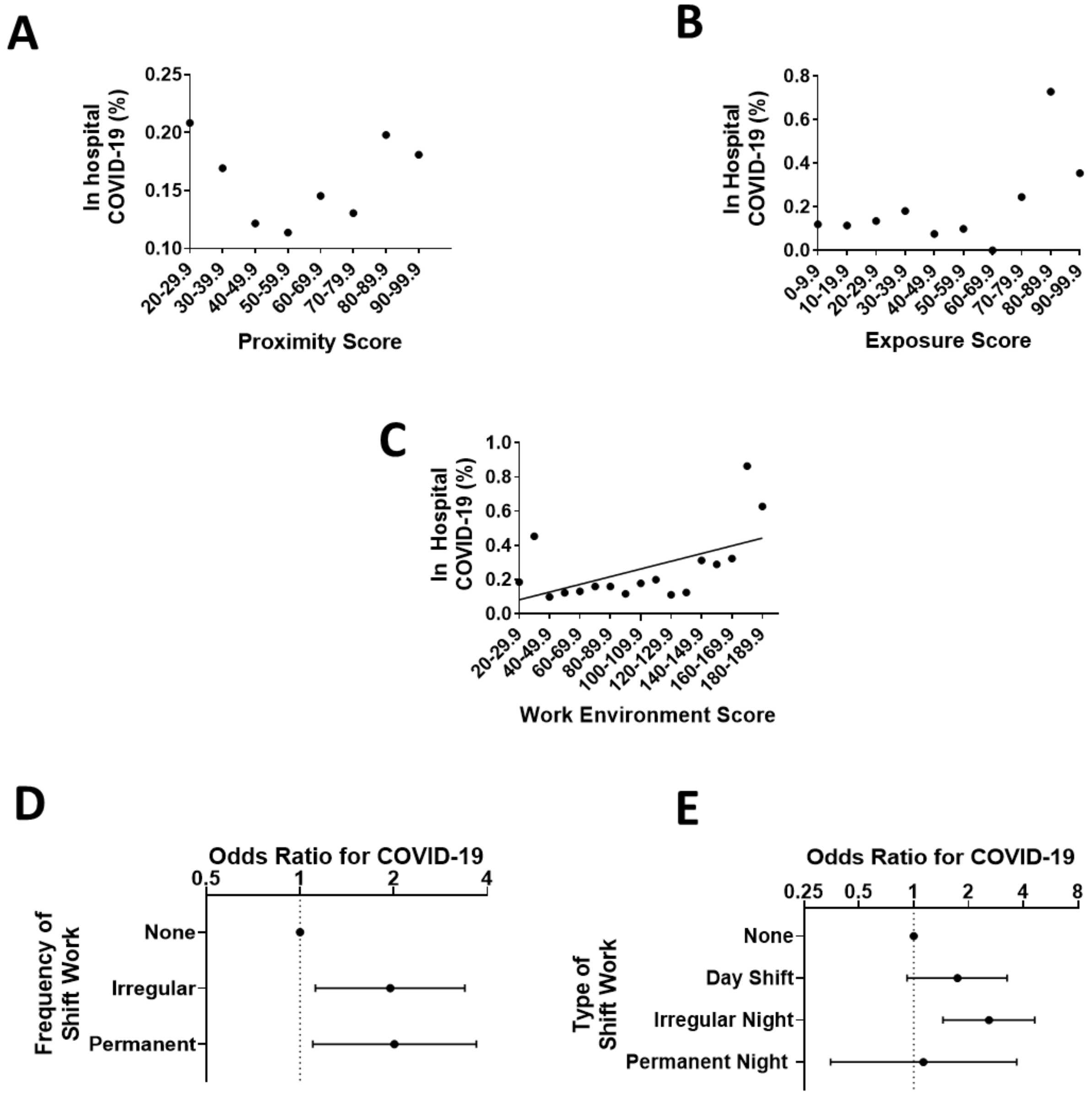


**Suppl. Figure 2: Effect of proximity and exposure through occupation on COVID-19 risk.** Occupational codes from the UK Biobank were combined with proximity and exposure data from ONS. **A** Risk of COVID-19 as a function of physical proximity. **B** Risk of COVID-19 risk as a function of exposure. **C** Risk of COVID-19 as a function of the sum of proximity and exposure scores (work environment score). **D** Shows the association of frequency of shift work with COVID-19 when the covariates in Model 3 and the work environment score are included in the model **E** Shows the association of the type of shift work with COVID-19 when the covariates in Model 3 and the work environment score are included in the model. Where significant linear regression lines are shown.

| | Reported Type of shift work | | | | |
|---|---|---|---|---|---|
| | None | Day Shift Workers | Irregular night shift work | Permanent night shift work | P values |
| N | 235135 | 24245 | 17971 | 7038 | |
| Age (years) | 52.9 (7.12) | 52.5 (7.08) | 51.11 (6.87) | 51.48 (6.9) | <0.01 |
| Sex (% male) | 46.61 | 47.68 | 62.76 | 61.72 | <0.01 |
| BMI (kg/m^2) | 27.09 (4.65) | 27.79 (4.99) | 28.22 (4.9) | 28.5 (4.87) | <0.01 |
| **Smoker (%)** | | | | | <0.01 |
| Never | 58.11 | 53.64 | 52.73 | 52 | |
| Previous | 31.89 | 32.08 | 30.64 | 30.02 | |
| Current | 9.75 | 13.92 | 16.15 | 17.68 | |
| Smoking pack-years | 19.99 (16) | 22.85 (17.49) | 24.23 (17.75) | 25.54 (18.32) | <0.01 |
| Daily alcohol intake (%) | 20.46 | 16.95 | 16 | 10.2 | <0.01 |
| Sleep Duration (h) | 7.05 (1.03) | 6.95 (1.22) | 6.85 (1.3) | 6.67 (1.51) | <0.01 |
| **Chronotype (%)** | | | | | <0.01 |
| Morning | 23.34 | 25.47 | 22.91 | 19.22 | |
| Evening | 8.01 | 7.88 | 9.84 | 16.93 | |
| **Ethnicity (%)** | | | | | <0.01 |
| White British | 88.5 | 83.35 | 80.05 | 81.05 | |
| White Other | 6.44 | 7.08 | 7.02 | 6.02 | |
| Mixed | 0.65 | 0.88 | 0.97 | 0.88 | |
| Asian | 1.71 | 3.57 | 3.8 | 3.42 | |
| Black | 1.39 | 2.66 | 4.86 | 5.36 | |
| Chinese | 0.34 | 0.49 | 0.45 | 0.67 | |
| Other | 0.69 | 1.61 | 2.42 | 2.26 | |
| Weekly work hours | 34.24 (13.19) | 34.98 (13.21) | 39.33 (14.55) | 39.62 (13.71) | <0.01 |
| Single Occupancy (%) | 15.63 | 18.76 | 18.72 | 18.49 | <0.01 |
| Urban area (%) | 85.98 | 89.56 | 89.33 | 90.97 | <0.01 |
| Townsend Index | -2.24 (-3.7 to 0.18) | -1.31 (-3.18 to 1.6) | -1.25 (-3.17 to 1.8) | -1.04 (-3.02 to 2.06) | <0.01 |
| High Cholesterol (%) | 7.88 | 8.57 | 8.55 | 9.29 | <0.01 |
| Diabetes (%) | 3.22 | 4.28 | 4.62 | 4.62 | <0.01 |
| Hypertension (%) | 20.33 | 22.24 | 22.06 | 23.26 | <0.01 |
| Depression (%) | 4.61 | 5.47 | 4.52 | 4.69 | <0.01 |
| Cardiovascular Disease (%) | 2.27 | 2.72 | 2.49 | 2.9 | <0.01 |
| Impaired Renal Function (%) | 0.09 | 0.14 | 0.08 | 0.09 | 0.15 |
| Defined Asthma (%) | 4.93 | 5.28 | 4.79 | 5.14 | 0.06 |
| COPD (%) | 0.13 | 0.23 | 0.19 | 0.13 | <0.01 |
| Liver Disease (%) | 0.53 | 0.49 | 0.58 | 0.5 | 0.57 |

**Suppl. Table 1: Demographics by type of shift work:** (n=284,389) Variables are expressed as mean (±SD) or as percentages. P values show whether there is a significant difference between groups (ANOVA).

| | Shift Work Type | | | | |
|---|---|---|---|---|---|
| | None | Day Shift Workers | Irregular Night Shift Workers | Permanent night shift workers | P values |
| N | 168,617 | 15,442 | 11,270 | 4,340 | |
| Proximity Score | 57.76 (13.1) | 62.5 (13.63) | 64.68 (13.93) | 64.69 (12.3) | <0.01 |
| Exposure Score | 17.31 (17.94) | 21.37 (19.31) | 24.16 (23.59) | 22.1 (20.52) | <0.01 |
| Work Environment Score | 75.07 (28.05) | 83.87 (28.37) | 88.83 (34.29) | 86.8 (30.12) | <0.01 |

**Suppl. Table 2: Work environment score by type of shift work:** (n=198,061) Variables are expressed as mean (±SD). P values show whether there is a significant difference between groups (ANOVA).
